# PARP1 regulates establishment of the ovarian reserve and age at natural menopause

**DOI:** 10.1101/2025.06.18.25329841

**Authors:** Hannah R. Schorle, Jason Halliwell, Jonas D. Elsborg, Stasa Stankovic, Emily Morbey, Jack Murzynowski, Amy V. Kaucher, Ruiqi Sun, Michael L. Nielsen, Kathleen Stewart-Morgan, Anna Murray, Jesper V. Olsen, John R. B. Perry, Eva R. Hoffmann

**Author notes:** Corresponding Author: Eva R. Hoffmann^1^.

## Abstract

Menopause not only signifies the end of a woman’s reproductive life, but also brings about significant changes in both mental and physical health due to hormonal changes. Age of natural menopause (ANM) and reproductive lifespan is determined by the formation and subsequent depletion of follicles in females. In this study, we identified a robust association between a common functional missense variant in *PARP1* and age at menopause in a sample of 201,323 women. Using an *in vitro* model to generate primordial germ cells - the precursors of sperm and oocytes- we report that *Parp1* affects PGCs in two different ways. Our identified missense variant (V762A) significantly reduces the germ cell population by enhanced DNA binding resulting in increased apoptosis. Conversely, deletion of Parp1 results in an increased population of PGCs and proteomic analysis revealed that transcription factors that drive PGC formation show increased expression in precursor cells. This is caused by increased transcription of *Rhox* genes (*Rhox2A*, *Rhox6* and *Rhox9*), implicating Parp1 in downregulation of genes that determine PGC fate. We propose that Parp1 regulates PGC fate and that its increased chromatin association (‘trapping’) causes a decreased population of PGCs, thereby providing a molecular cause of the earlier onset menopause on female carriers of the variant.

## Introduction

Menopause marks the end of a women’s reproductive lifespan and occurs naturally between the ages of 45 and 55 years (Schoenaker et al., 2014). The age at natural menopause (ANM) varies regionally across the world and is clinically defined as the cessation of menstruation for 12 consecutive month and accompanied by a decline in oestradiol and an increase of luteinizing hormone and follicle stimulating hormone (Yang et al., 2024). The timing of menopause profoundly impacts women’s health, as all-cause mortality is reduced by 2% with each increasing year of ANM (Ossewaarde et al., 2005, Mondul et al., 2005). An earlier ANM is associated with decreased risk of breast (Kelsey et al., 1993) and ovarian (Schettler, 1976) cancers, but increased risk of cardiovascular disease (Atsma et al., 2006), stroke (Lisabeth et al., 2009) and osteoporosis (Gallagher, 2007), while a later onset of ANM increases the risk for hormone-sensitive cancers (Cancer, 2019, Day et al., 2015, Ruth et al., 2021).

ANM is determined by the formation and depletion of follicles in the ovary, consisting of germ cells and surrounding somatic cells. Primordial germ cells arise early during embryo development and eventually give rise to the finite follicle pool each women is born with (reviewed in Findlay et al., 2015). The heritability of ANM is estimated to be around 50% and additive models reveal effect sizes of common and rare variants that range from a few weeks to 5.1 years (Ruth et al., 2021, Murabito et al., 2005). About 2 out of 3 genetic variants have functions in highly conserved DNA damage response and repair pathways, and include variants that, when homozygous or hemizygous, result in early cessation of reproductive lifespan, referred to as premature ovarian insufficiency (POI; ANM before 40 years) (Ruth et al., 2021, Stankovic et al., 2024).

PARP proteins use nicotinamide adenine dinucleotide (NAD^+^) to synthesize poly(ADP-ribose; referred to as PAR) and catalyze their addition to side chains of target proteins, including themselves in response to DNA damage (Lüscher et al., 2022, Chambon et al., 1963, Ray Chaudhuri and Nussenzweig, 2017). However, PARP proteins of which there are at least 18, also have functions outside of DNA repair including in chromatin remodeling and transcriptional regulation (Gill et al., 2022, Huletsky et al., 1989, Zong et al., 2022, Huambachano et al., 2011). PARP1, the most abundant protein of the PARP family, acts as a sensor of single- and double-stranded DNA breaks and facilitating repair by recruiting key factors such as XRCC1 in the base-excision repair pathway through PARylation activity, which in turn releases PARP1 from sites of DNA damage ensuring efficient repair processes (Caldecott et al., 1994, Caron et al., 2019, Demin et al., 2021). The release of PARP1 from damaged DNA is a crucial step, and ‘trapping’ of PARP1 to DNA has been shown to be cytotoxic, a process that has been taken advantage of in cancer treatment (Murai et al., 2012, Zandarashvili et al., 2020). Over the years, additional roles of PARP1 have been uncovered, including chromatin conformation, transcriptional regulation and the re-initiation of stalled replication forks (Maya-Mendoza et al., 2018, Zong et al., 2022, Gill et al., 2022, Hassa and Hottiger, 2002, Cohen-Armon et al., 2007). Physiologically, PARP1 is involved in many processes such as carcinogenesis, ageing, inflammation and neuronal function (reviewed in Kim et al., 2005).

PARP1 is part of the base excision repair (BER) pathway, which plays a key roles in DNA demethylation during primordial germ cell (PGC) development (Hajkova et al., 2010), when the genome-wide methylation marks are erased. Active DNA demethylation in PGCs involves the conversion of 5-methylcytosine (5mC) to 5- hydroxymethylcytosine (5hmC) by TET enzymes, followed by further oxidation and excision through BER (Hajkova et al., 2010, He et al., 2011, Tahiliani et al., 2009). PARP1 facilitates this process by recruiting BER factors, including XRCC1, which acts as a scaffold for DNA repair proteins such as DNA ligase III. This mechanism ensures efficient replacement of modified cytosines, contributing to epigenetic reprogramming in germline development (Hajkova et al., 2010).

There are limited studies exploring the role of PARP1 in oocytes and germ cells, with most of the available research focusing on the adult ovary (Makogon et al., 2010, Nakamura et al., 2020, Qian et al., 2010). Exposure to the Parp1/2 inhibitor olaparib, reduced follicle numbers in *in vitro* cultured mouse ovaries, lowered oocyte retrieval rates, and impaired fertilization success, likely due to granulosa cell dysfunction (Nakamura et al., 2020). In *Parp1^-/-^* mice, fetal oocytes displayed meiotic defects such as incomplete homologous chromosome synapsis and persistent DNA damage as well as chromatin defects in adult oocytes. Despite these phenotypes, litter size and frequency was reportedly not affected (Yang et al., 2009). Thus, *Parp1^-/-^* female mice are healthy and fertile (Yang et al., 2009, Wang et al., 1995).

DNA repair pathways and checkpoint signaling proteins, such as CHEK1 and CHEK2 identified in human genomic studies are supported by *in vivo* studies of mouse models that show a reduced or extended reproductive lifespan (Ruth et al., 2021). Although mouse models are an attractive means to assess the function of candidate variants identified in human genomic studies, it is often challenging to study the earliest stages of gametogenesis in mice, as the founding germ cells arise during the early stages of embryogenesis when there are only a small number of cells in the embryo (Ohinata et al., 2009). Recent developments in recapitulation of early germ cell development *in vitro* from mouse embryonic stem cells, provide new opportunities for mechanistic insight into gene and variant function throughout gametogenesis (Hayashi et al., 2011a).

In this study, we utilize large-scale population data to demonstrate that alleles in PARP1, including *V762A*, influence menopause timing in the general population. We further elucidate the mechanistic role of *Parp1* in regulating early germ cell development *in vitro*, using a widely used model to generate primordial germ cell-like cells (PGCLC) starting from mouse embryonic stem cells (Hayashi et al., 2011a, Hayashi et al., 2012). *Parp1^V762A/V762A^* variant mESCs show reduced efficiency in differentiating into PGCLCs, with higher Parp1-V762A DNA binding and increased apoptosis. In contrast, *Parp1^-/-^* cells display an enhanced PGCLC population, with no changes in proliferation or apoptosis. RNA and proteomic analyses reveal that *Rhox* genes, key to germ cell development (Li et al., 2023a, Berletch et al., 2013, Daggag et al., 2008), are differentially expressed in *Parp1^-/-^*cells. We infer that PARP1 likely represses *Rhox* genes, which are critical regulators of early germ cell differentiation.

## Results

### A functional missense variant in PARP1 is associated with age at natural menopause

A previous large-scale population study identifying germline genetic determinants of mosaic Y chromosome loss (LOY) in blood – a biomarker of DNA damage response – identified a robust genome-wide significant association between coding variants in PARP1 and LOY (Thompson et al., 2019). The identified missense variant (V762A) had a minor allele (alanine substitution) which is protective against LOY and has experimentally been shown to reduce the catalytic activity of PARP1 by 30–40% (Zaremba et al., 2011). We hypothesized that this variant would provide a genetic instrument which would allow us to model the effect of PARP inhibition on age at menopause in women. Using published genome-wide association study (GWAS) data in 201,323 women of European ancestry, we found that the PARP inhibiting allele was suggestively associated with earlier age at women (rs1136410, effect: -0.075 years per allele [95% CI: 0.04-0.11], P=4.6×10^-6^).

We sought to complement this analysis by interrogating the role of rare (<0.1% minor allele frequency) protein-coding variants captured through exome-sequencing in the UK Biobank study. Using results generated in 106,973 women of European ancestry (Stankovic et al., 2024), we identified 41 women carrying heterozygous protein truncating variants in PARP1 and a further 353 women carrying predicted deleterious missense variants (defined as CADD >= 25). Whilst we saw no significant association with menopause timing in the PTV carriers (beta=-0.28 years, P=0.49), we observed a nominally significant effect of earlier menopause in the predicted deleterious missense variant carriers (beta= -0.69 years, P=9.6×10^-3^). Notably this effect is almost an order of magnitude larger than the more common V762A missense variant which was not included in this analysis. Collectively, these data suggest that functional variants in the *PARP1* gene influence menopause timing in the general population.

### Generation of *Parp1^-/-^, Parp1^V762A/V762A^* and *XRCC1^-/-^* mouse embryonic stem cells

Our genetic analyses demonstrate that women carrying the *Parp1^V762A/V762A^* variant experience an earlier onset of menopause. However, the precise mechanism and the specific stage of germ cell development affected by this variant, or indeed PARP1 itself, remain unclear. Primordial germ cells (PGCs) are the precursors to both oocytes and spermatocytes, and form early during embryonic development, around embryonic day 6.25 in mouse (Chiquoine, 1954). In females, the initial number of PGCs determines the number of primordial follicles in the ovarian reserve, which is a critical determinant of reproductive lifespan (Depmann et al., 2015). PARP1 is active in the base excision repair pathway and catalyzes the formation of PAR chains at DNA break (Hajkova et al., 2010, Caron et al., 2019). We therefore explored the effect of *Parp1^V762A/V762A^*, which affects the catalytic domain of PARP1 (Wang et al., 2007), on PGC formation in vitro (Hayashi et al., 2011a).

*Parp1* is highly conserved between humans and mice (Schreiber et al., 2006) and to further analyze the amino acid residue at position 762, we aligned human and mouse PARP1 protein sequences using T-Coffee, a multiple sequence alignment tool (Notredame et al., 2000). The region surrounding the 762 amino acid residue was found to be highly similar (Figure 1A).

**Figure 1:**
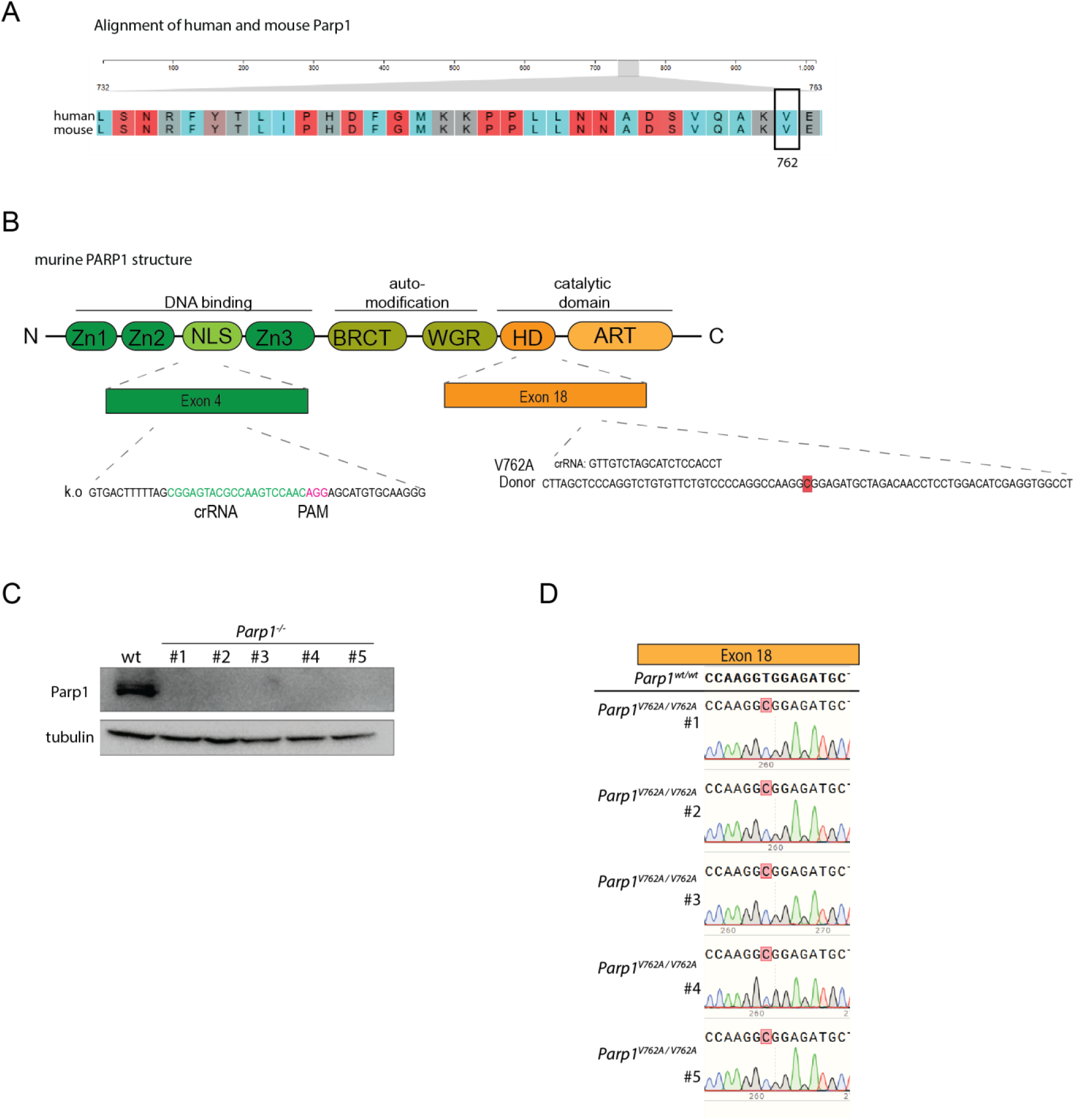
Generation of *Parp1* mESC clones using gene editing. **(A)** Amino acid alignment of human and mouse PARP1 proteins around the V762A amino acid residue, highlighted in a black box. **(B)** Structural overview of the murine PARP1 protein. The guide RNA target sites used for generating PARP1- knockout cells and introducing the V762A variant are shown on exon 4 and exon 18. **(C)** Western blot showing bands forPARP1 (113kDa) and tubulin (55kDa) protein in *Parp1^wt/wt^* and *Parp1^−/−^* mESCs. Successful deletion of *Parp1* gene indicated by the absence of expected band. Five *Parp1^−/−^* clones were analyzed and subsequently used in this study. **(D)** DNA sequences from sanger sequencing displaying validation of the *Parp1^V762A/V762A^* variant in mESCs. The wild-type sequence (*Parp1^wt/wt^*) is shown at the top, with the variant resulting from a T>C substitution. Five independent *Parp1^V762A/V762A^* clones were chosen and used in this study.

To generate *Parp1^-/-^, Parp1^V762A/V762A^*, and *Xrcc1^-/-^* mESC clones, we employed a CRISPR-Cas9 gene editing approach (Cong et al., 2013). For the knockout, we designed guides targeting the N-terminal domains of the proteins (Exon 4 for Parp1 and Exon 2 for *Xrcc1*; Figure1B; Figure S1A). To generate a mESC clone with the *Parp1^V762A/V762A^* variant, we designed a donor oligo targeting the helical domain (HD) of the Parp1 catalytic region resulting in a T>C base pair change that leads to an amino acid substitution from valine to alanine (Figure 1B). Lastly, clones that were subjected to CRISPR editing, but showed no evidence of editing, were selected as the *Parp1^wt/wt^* control group. Using this strategy we generated five *Parp1^-/-^* clones, five *Parp1^V762A/V762A^* clones and five *Xrcc1^-/-^* clones that were analyzed independently for each experiment, unless otherwise stated. The knock-out of *Parp1* and *Xrcc1* were confirmed via western blot (Figure 1C), and introduction of the *Parp1^V762A/V762A^*variant was verified using Sanger sequencing (Figure 1D).

### *Parp1^V762A/V762A^* reduces the PGCLC population *in vitro*

We utilized a previously published protocol to generate primordial germ cell-like cells (PGCLCs) *in vitro* to understand the role of PARP1 in primordial germ cell formation (Hayashi et al., 2012). This process involves a two-step differentiation of mESCs into Epiblast-like cells (EpiLCs) and subsequently into PGCLCs. Day 2 EpiLCs are cultured in low binding U-bottom plates, allowing for the aggregation of the cells. Within the aggregates, termed embryoid bodies (EBs) the PGCLCs form. The subpopulation of PGCLC within each embryoid bodies can be sorted and analyzed using fluorescent activated cell sorting (FACS) (Figure 2A). The mESC lines used in this study harbor *Blimp1-Venus* and *Stella-ECFP* reporters, both genes are markers for germ cell development (Magnúsdóttir et al., 2013, Hackett et al., 2018). In mouse primordial germ cells *Blimp1* is expressed around E6.25 and together with *Prdm14* and *Tfap2C* is essential for PGC specification (Magnúsdóttir et al., 2013). *Prdm14* induces the expression of Stella or *Dppa3* later at E7.5 (Hackett et al., 2018). Using these markers, we are able to detect early (*Blimp1-Venus*) and later (*Stella-ECFP*) primordial germ cells. We compared the *in vitro* derived PGCLC population (Blimp1-Venus positive) on day 6 of differentiation between *Parp1^wt/wt^*and *Parp1^V762A/V762A^* samples via fluorescent activated cell sorting (FACS) (Figure 2B). We observed a significant reduction of the Blimp1-Venus positive population in the *Parp1^V762A/V762A^* compared to *Parp1^wt/wt^* (Figure 2B; d = -0.83; p = 3.2*10^-3^, Wilcoxon rank-sum test). This suggests that the PARP1 variant reduces the PGCLC population *in vitro*.

**Figure 2:**
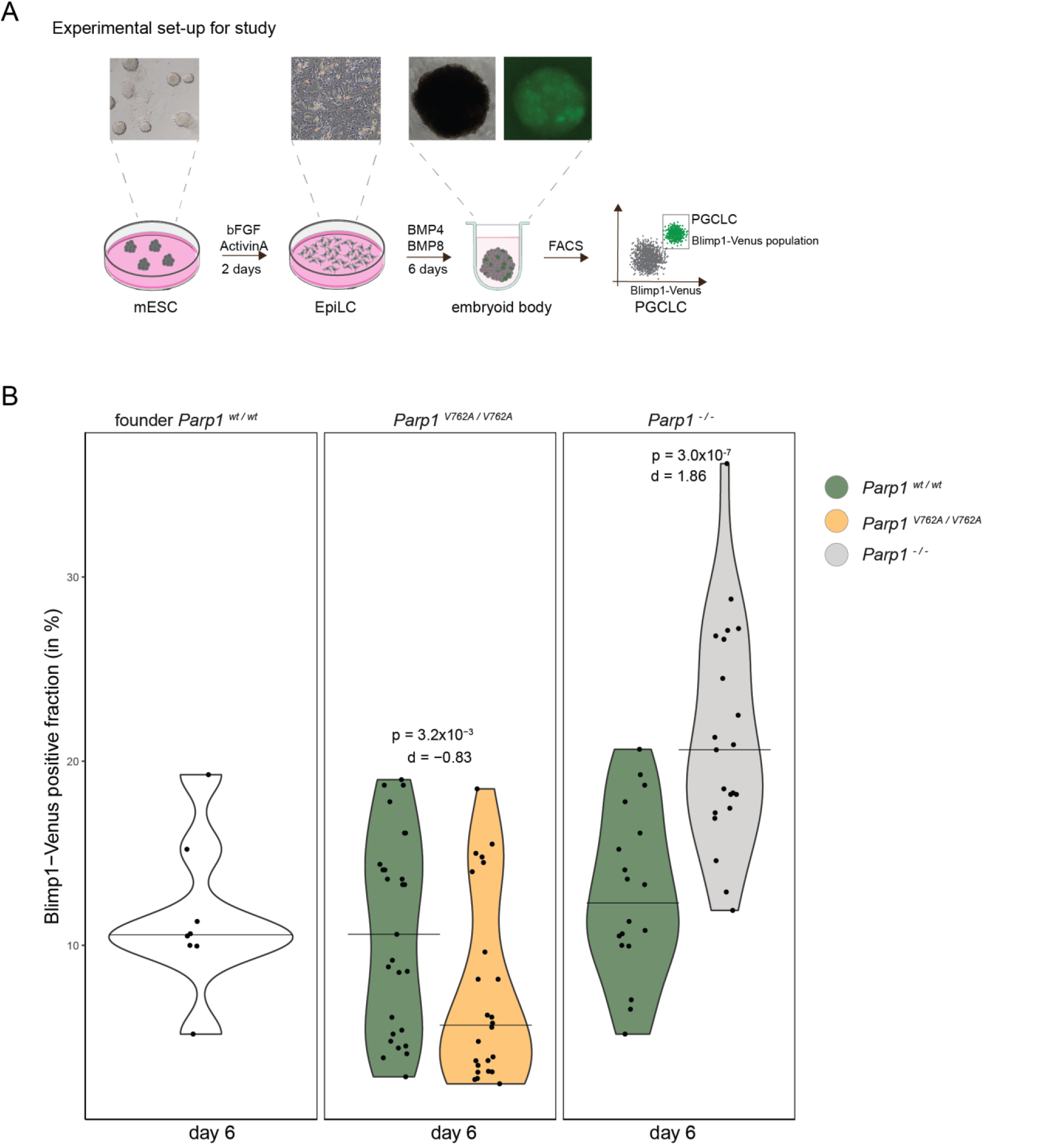
Generation of primordial germ cell like cells (PGCLC) from *Parp1* clones shows reduced PGCLCs in the variant, but increased PGCLCs in *Parp1^wt/wt^*. **(A)** Schematic overview of the experimental workflow for generating primordial germ cell-like cells (PGCLCs). Mouse embryonic stem cells (mESCs) were differentiated into epiblast-like cells (EpiLCs) using bFGF and Activin A over two days. On day 2, EpiLCs were transferred to low-attachment U-bottom wells and cultured with BMP4 and BMP8, promoting aggregation and subsequent formation of embryoid bodies (aggregates) containing PGCLCs. Fluorescent-activated cell sorting (FACS) was used to isolate Blimp1-Venus-positive PGCLCs on days 2, 4, and 6. **(B)** Violin plot displaying the Blimp1-Venus positive cells in percent (%) on day 6, for the founder (not CRISPRed cells), the Parp1^wt/wt^ that did not show Parp1 deletion after CRISPR as a control (Parp1^wt/wt^), the variant Parp1^V762A/V762A^ and Parp1^-/-^ quantified via FACS. Statistical significance was assessed using the non- parametric Wilcoxon test with Bonferroni adjustment. Effect sizes (Cohen’s d) are indicated.

To determine whether the effect seen during PGC differentiation was caused by the *Parp1* genotype or other experimental variables, we performed ANOVA analysis, where we tested the influence of clonal variance, experimental replicates and *Parp1* genotype status (Table 3). The analysis revealed that the genotype had a significant effect on PGCLC differentiation (F= 42.58, p = 1.39×10⁻⁷).

### **Parp1^V762A/V762A^** traps PARP1 to the DNA, lowers NAD^+^ levels in mESCs and increases apoptosis in PGCLCs

Previous studies found that PARP1^V762A/V762A^ exhibits a reduced enzymatic activity when compared to wild-type PARP1 (Wang et al., 2007). We were interested in whether the variant would also impact PARP1 ‘trapping’, the increased association with and/or retention of PARP1 on the DNA. *Xrcc1^-/-^* is known to increase PARP1 trapping on chromatin (Demin et al., 2021). Thus, we isolated the chromatin fraction from mESCs and measured the amount of DNA-bound PARP1 (Gatti et al., 2020) in *Parp1^V762A/V762A^* compared to *Parp1^wt/wt^*cells (Figure 3A). As a positive control, we included three *Xrcc1^-/-^* clones, where a larger proportion of PARP1 association with chromatin is expected (Demin et al., 2021). Using tubulin as a loading control we measured the relative amount of PARP1 in the chromatin bound fraction. However, we found a higher amount and more variability of chromatin bound PARP1^V762A/V762A^ when compared to the *Parp1^wt/wt^* (Figure 3A). Consistent with previous findings, the *Xrcc1^-/-^* clones also displayed elevated levels of PARP1 bound to the DNA (Figure 3A). We infer that the *Parp1^V762A/V762A^* variant increases PARP1 trapping either through prolonged chromatin retention or a higher amount of PARP1^V762A/V762A^ on the DNA.

**Figure 3:**
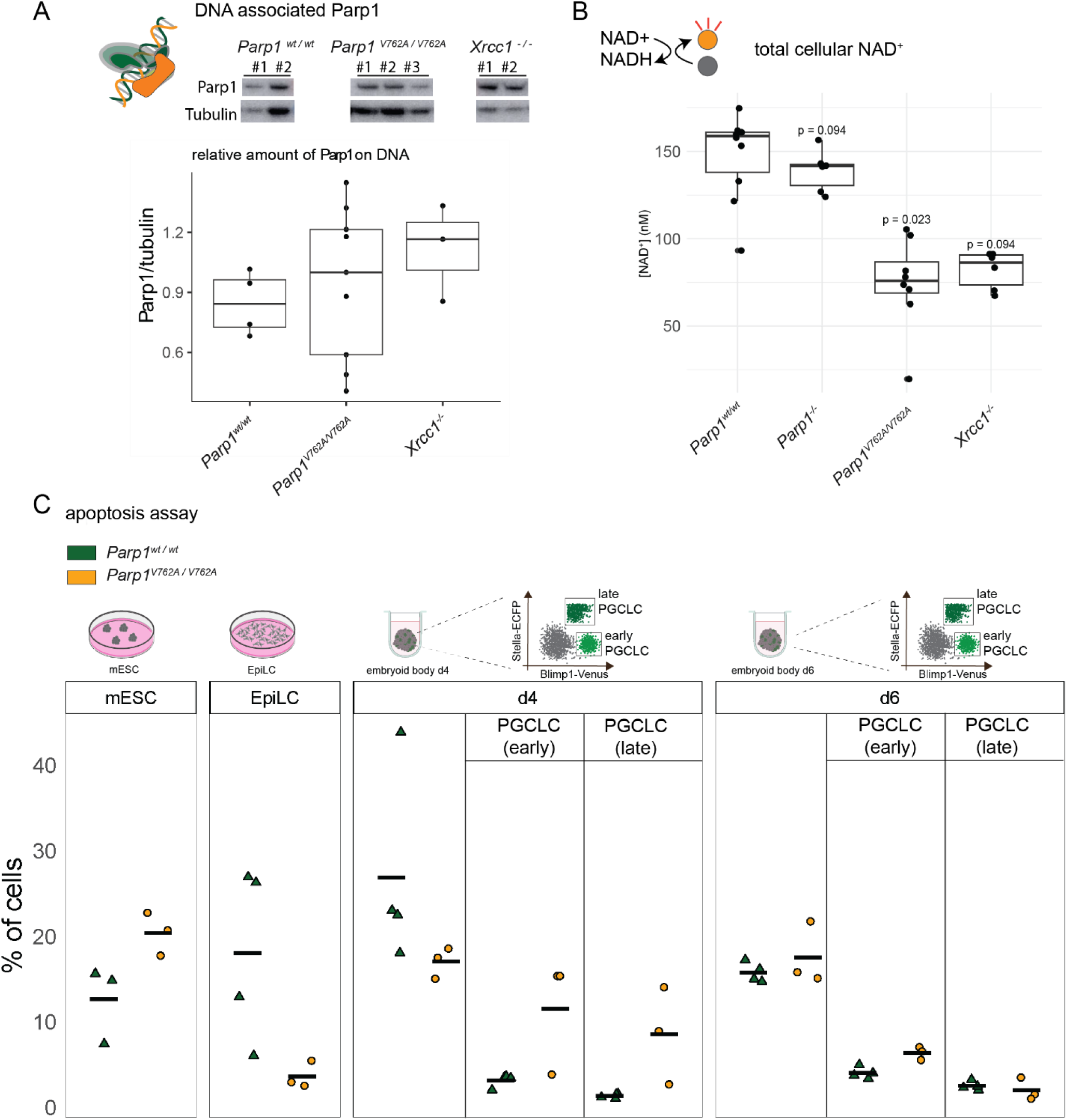
Functional analysis of *Parp1^V762A/V762A^* mESCs shows elevated trapping and cell death. **(A)** Boxplot of chromatin-bound PARP1 in *Parp1^wt/wt^, Parp1^-/-^, Xrcc1 ^-/-^,* and *Parp1^V762A/V762A^* mESCs. Chromatin fractions were isolated and analyzed via SDS-PAGE, with chromatin-bound PARP1 quantified relative to tubulin. Representative western blot images of PARP1 and tubulin are shown. Analysis of band intensity through ImageJ. Statistical analysis: non-parametric Wilcoxon test with Bonferroni correction. **(B)** Boxplot of total cellular NAD⁺ levels measured in *Parp1^wt/wt^ ^t^*, *Parp1^-/-^, Xrcc1^-/-^,* and *Parp1^V762A/V762A^* mESCs using a colorimetric assay. Statistical analysis: non-parametric Wilcoxon test with Bonferroni correction. **(C)** Boxplots depicting apoptosis analysis via AnnexinV and PI of *Parp1^wt/wt^* and *Parp1^V762A/V762A^* cells across differentiation stages (mESCs, EpiLCs, and aggregates/embryoid bodies with PGCLCs on days 4 and 6). Early and late germ cell populations were distinguished using Blimp1-Venus and Stella-ECFP markers. Statistical analysis: non-parametric Wilcoxon test.

The binding of PARP1 to DNA activates its enzymatic function, triggering PARylation (Dawicki-McKenna et al., 2015). In this process PARP1 uses NAD^+^ as a substrate in generating poly(ADP-ribose) chains, tagging itself and other proteins (Kim et al., 2005). As we observed a high amount of trapped PARP1, we were interested in seeing the effect of *Parp1^V762A/V762A^* and quantified total cellular NAD^+^ levels using a colorimetric assay, which detects the reduction of NAD^+^ to NADH and its subsequent oxidation (Figure 3B). We found a significantly lower total cellular NAD^+^ in the *Parp1^V762A/V762A^* than in the *Parp1^wt/wt^* (d = -2.88; p= 0.023, Wilcoxon rank-sum test; Figure 3B). The *Parp1^-/-^* and *Xrcc1^-/-^* mESC cell lines showed equivocal levels of NAD^+^ compared to the wild-type cell lines (d= -0.42; p = 0.094, Wilcoxon test for *Parp1^-/-^* and d= -3.17; p=0.094, Wilcoxon test). We infer that prolonged retention or increased chromatin association of PARP1-V762A results in a greater NAD^+^ consumption compared to the wild-type protein.

PARP1 trapping results in cellular toxicity and subsequent apoptosis (Demin et al., 2021, Murai et al., 2012, Krastev et al., 2022). To investigate the impact of the PARP1^V762A/V762A^ on cell death, we performed an apoptosis assay by staining for Annexin V and propidium iodide that mark early apoptotic (Annexin V positive, PI negative) and late apoptotic cells (Annexin V positive, PI positive; Figure 3C; Supplementary Fig. S2A: schematic overview). We then analysed the percentage of apoptotic cells (AnnexinV positive and AnnexinV plus PI double positive) in mESCs, EpiLCs and day 4 and 6 embryoid bodies and gated PGCLCs on day 4 and day 6 using Blimp- Venus and Stella-ECFP (Figure 3C). We found a higher percentage of mESCs in apoptosis in *Parp1^V762A/V762A^*when compared to *Parp1^wt/wt^*, but a higher amount of apoptotic cells in *Parp1^wt/wt^* EpiLCs (Figure 3C). The apoptotic fraction on day 4 and day 6 in the embryoid bodies were similar. To assess any potential differences in apoptosis in PGCLCs, we gated for Blimp1-Venus positive and Stella-ECFP negative (early PGCLCs), as well as Blimp1-Venus positive and Stella-ECFP positive (late PGCLCs) on day 4 and day 6 (Figure 3C). We were able to detect a higher amount of apoptotic cells in the *Parp1^V762A/V762A^* on day 4 in early as well as late PGCLCs. The fraction of apoptotic PGCLCs on day 6 was similar between the *Parp1^V762A/V762A^* and the *Parp1^wt/wt^*.

Together we find that the *Parp1^V762A/V762A^* cells exhibit a lower total cellular NAD^+^ concentration, an increased amount of DNA-associated PARP1, and a higher fraction of apoptotic cells during PGCLC differentiation. This suggests that the *Parp1^V762A/V762A^* variant displays a higher degree of trapping on the DNA, continues to carry out PARylation, and thereby decreases the total cellular NAD^+^. Consequently, the trapped PARP1 is cytotoxic and leads to apoptosis of PGCLCs.

### Parp1 knockout increases the PGCLC population without affecting proliferation or apoptosis

To further study the impact of Parp1 activity on germ cell development, we used our *Parp1^-/-^* mESCs to generate PGCLCs. In contrast to the cell line carrying the *Parp1^V762A/V762A^* variant, *Parp1^-/-^* mESCs exhibited a higher population of cells expressing Blimp1-Venus compared to control (*Parp1^wt/wt^*) upon induction of PGCLC differentiation (Figure 2B; d=1.86; p=3×10^-7^; Wilcoxon test). We infer that PARP1 has a suppressive role in PGCLC differentiation.

Given the significant upregulation of the PGCLC population observed on day 6 in the *Parp1^-/-^* compared to the control *Parp1^wt/wt^* clones, we extended our analysis to earlier stages (day 2 and day 4) of the differentiation protocol to investigate the temporal dynamics of *Blimp1-Venus* expression (Figure 4A). In line with our findings from day 6 we also observed a significant increase in the Blimp1-Venus positive population at day 2 and day 4 in the *Parp1^-/-^* compared to the *Parp1^wt/wt^* clones (Figure 4A; d = 1.15, p=3.9×10^-3^ andd = 1.88, p = 2.5*10^-5^, respectively, for day 2 and 4; Wilcoxon test). To explore whether this is due to PARP1’s function in base excision repair, we generated an *Xrcc1^-/-^* mESC line and differentiated the cells to PGCLC (Figure S1A-C). The *Xrcc1^-/-^* clones showed significantly more variability but a lower average population of cells expressing Blimp- 1-Venus after 6 days of differentiation compared to clones that retained the wild-type *Parp1* gene. We conclude that PARP1 suppresses PGCLC differentiation in mESCs independently of its role in XRCC1 dependent DNA damage repair.

**Figure 4:**
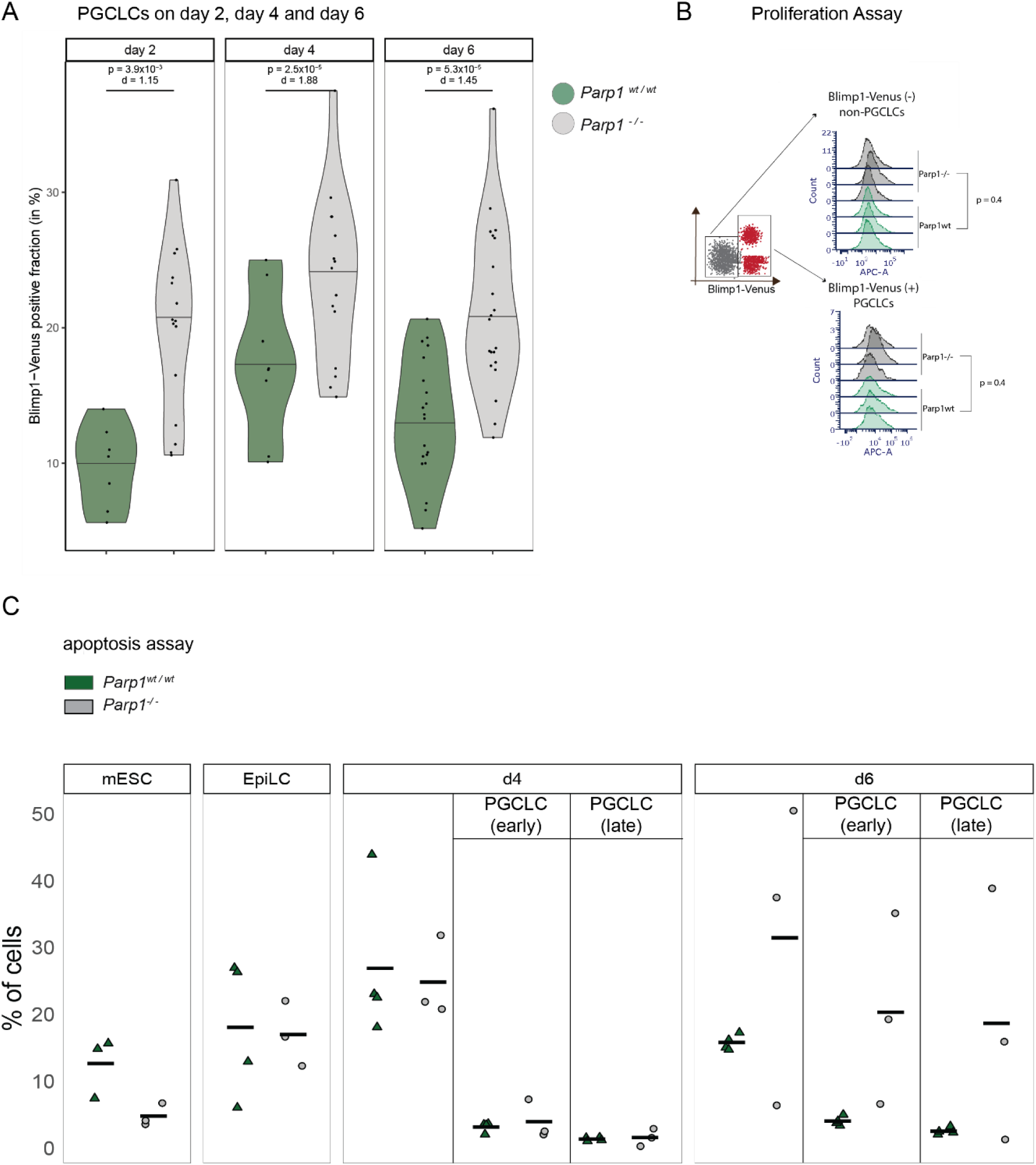
*Parp1* deletion enhances PGCLC populations across different stages without affecting proliferation or reducing apoptosis. **(A)** Violin plot displaying Blimp1-Venus positive PGCLC population in percentage (%) of *Parp1^-/-^* and *Parp1^wt/wt^* cell on day 2, day 4 and day 6 quantified through FACS. Statistical analysis: non-parametric Wilcoxon test (Bonferroni adjustment), effect size: Cohen’s d. **(B)** Histogram showing the proliferation assay comparing *Parp1^wt/wt^* (green) and *Parp1^-/-^* (black/gray) cells in day 6 PGCLCs and non-PGCLCs using CellTrace dye. Three independent *Parp1^wt/wt^* and *Parp1^-/-^* clones were analyzed. Statistical analysis: non-parametric Wilcoxon-test. Schematics FACS plot shown on the left depicting gating strategy. **(C)** Boxplots depicting apoptosis analysis from n=3 clones per condition of *Parp1^wt/wt^* and *Parp1^-/-^* cells across differentiation stages (mESCs, EpiLCs, and embryoid bodies containing PGCLCs on days 4 and 6) using Annexin V and PI labeling with FACS. Early and late germ cell populations were distinguished using Blimp1- Venus and Stella-ECFP markers. Statistical analysis: non-parametric Wilcoxon test.

To gain deeper insights into the molecular characteristics of these cells, we performed qPCR expression analysis of key stem cell and germ cell markers, across distinct stages of PGCLC differentiation. *Blimp1, Prdm14* and *Tfap2c* are expressed early during germ cell development and have been shown to be sufficient to induce PGC-like fate, by re-enforcing the transcription of the pluripotency markers *Nanog, Pou5f1* (OCT4) and *Sox2* while also repressing mesodermal genes (Magnúsdóttir et al., 2013). *Dppa3* (Stella) is expressed later, induced by PRDM14 (Hackett et al., 2018). We analyzed the expression of the stem cell markers *Oct4, Sox2* and *Nanog* and germ cell markers *Blimp1, Prdm14, Tfap2C*, *Stella* in mESCs, EpiLCs and FACS sorted PGCLCs on different days of differentiation (day 2, day 4 and day 6; Figure S2B, C). We did not detect any significant differences in the expression of any PGC or stem cell markers. This revealed that although a loss of PARP1 increased the proportion of PGCLCs, the cells retained similar germ cell identity by gene expression analysis.

A larger fraction of PGCLC in the *Parp1^-/-^* compared to the *Parp1^wt/wt^* could be due to upregulated proliferation, reduced cell death, or increased specification of PGCLC fate in the *Parp1^-/-^*. We first assessed proliferation using the CellTrace-APC dye, where cells a given a pulse of the dye, which subsequently is diluted in each cell division (Figure 4B). Thus, the reduction in intensity is correlated with the number of cell divisions (Tempany et al., 2018). We labelled day 2 EpiLCs with CellTrace-APC dye and subsequently analyzed on day 6 of PGCLC differentiation for the PGCLC (Blimp1-Venus positive) and non-PGCLC (Blimp1-Venus negative) population (Figure 4B). No significant differences in proliferation between the *Parp1^wt/wt^* and *Parp1^-/-^* were observed in either the PGCLC or non-PGCLC populations (d = 1.26 for non-PGCLC and d = 1.11 for PGCLC; p = 0.4, Wilcoxon test; Figure 4B). Thus, differences in proliferation between *Parp1^wt/wt^* and *Parp1^-/-^*cells are unlikely to contribution to the observed increased population of PGCLCs in the *Parp1^-/-^* clones.

Apoptosis was not reduced in the *Parp1^-^*^/-^ clones, as measured by the AnnexinV and PI fraction of mESCs, EpiLCs and PGCLCs. Instead, we detected an elevation of apoptosis on day 6 in the early (Blimp1-Venus positive and Stella-ECFP negative) and late PGCLCs (Blimp1-Venus positive and Stella-ECFP negative) of the *Parp1^-^*^/-^ cells (Figure 4C). Taken together, our findings suggest that *Parp1^-/-^* cells exhibit enhanced capacity for inducing PGCLC differentiation, but undergo cell death following the initial stages of germ cell initialization.

### Proteomics analysis reveals potential network leading to upregulated PGCLC population

Since elevated proliferation or reduced apoptosis cannot explain the increased fraction of PGCLCs in *Parp1^-/-^* compared to the *Parp1^wt/wt^*, we next asked whether PARP1 might have a suppressive function on the expression of genes that govern PGCLC differentiation. To address this, we compared the proteomes of mESC *Parp1^wt/wt^* and *Parp1^-/-^* by mass-spectrometry. Interestingly, most proteins are not regulated by PARP1 knockout. However, we were able to quantify two proteins which are differentially expressed between *Parp1^wt/wt^* and *Parp1^-/-^* conditions (Figure 5A) including PARP1, which, as expected, was absent in the *Parp1^-/-^* (Figure 5A, B). Conversely, we found RHOX2a/e expression to be significantly elevated in cells with *Parp1^-/-^*compared to *Parp1^wt/wt^* (Figure 5A, C). *Rhox2a* belongs to the reproductive homeobox genes located on the X chromosome (Rhox) that are expressed in male and female reproductive tissue (Maclean et al., 2005). *Rhox2a,* specifically, has been detected in fetal testis and ovaries (Daggag et al., 2008). Our findings suggest that PARP1 downregulates *Rhox2a/e* expression, which could explain our observation that knockout cells preferentially differentiate into PGCLCs.

**Figure 5:**
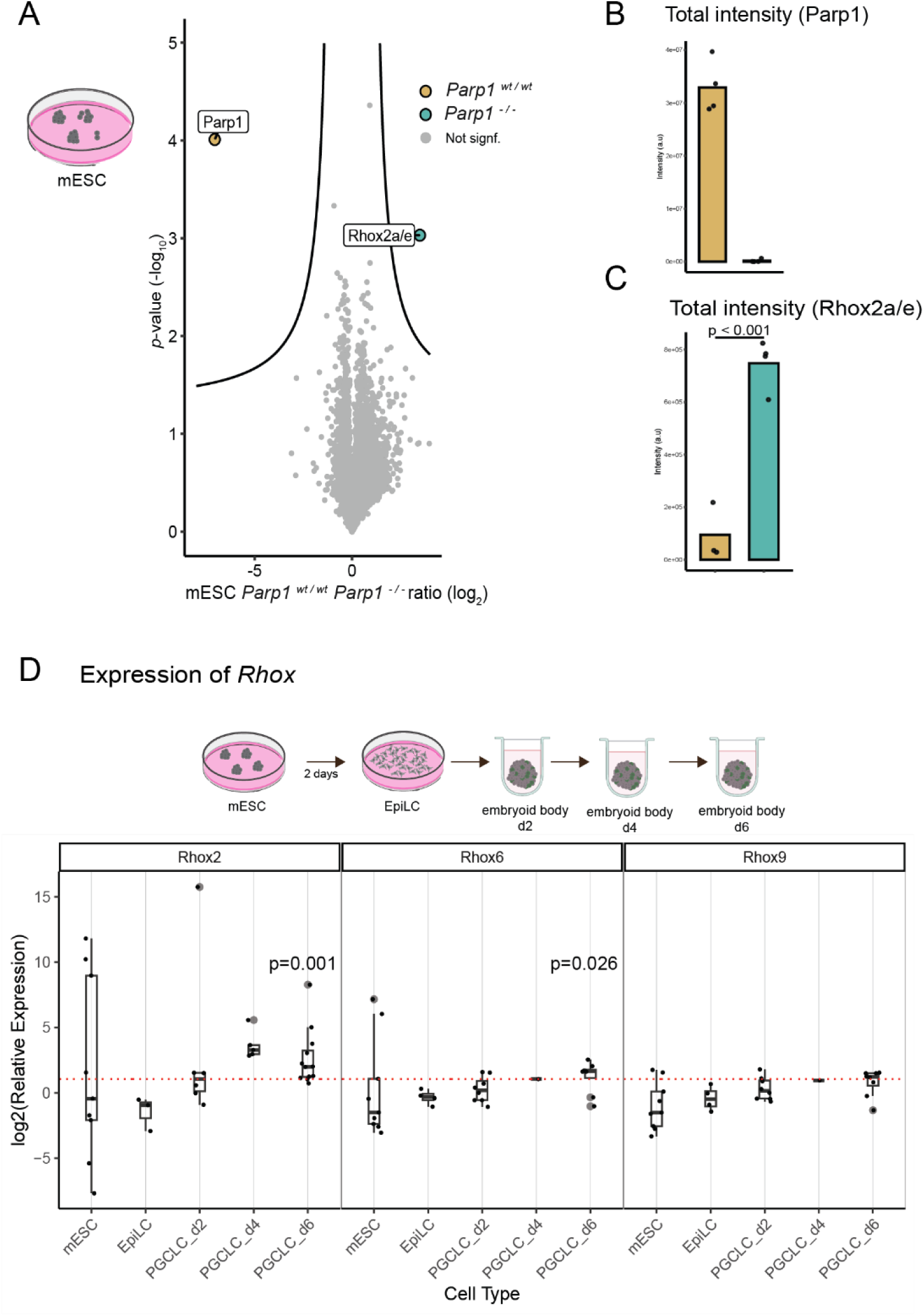
Proteomics and gene expression analysis in *Parp1^-/-^* cells reveals potential link to *Rhox* genes. **(A)** Volcano plot highlighting candidate proteins from proteomics analysis, that are significantly upregulated or downregulated in *Parp1^-/-^* cells compared to *Parp1^wt/wt^* cells. Four *Parp1^wt/wt^* and four *Parp1^-/-^* mESC clones were used in this analysis. **(B)** Barplot of relative intensity of PARP1 **(C)** Barplot relative intensity of RHOX2a/e (**D)** Boxplot displaying relative expression of potential candidate genes *Rhox6* and *Rhox9* by qPCR. Gene expression levels are presented using the delta-delta Ct (ΔΔCt) method, normalized to a reference gene (*Gapdh*) and relative to wild-type expression levels, each data point shown as a dot. Statistical analysis: non-parametric Wilcoxon test

Both *Rhox6* and *Rhox9* are highly expressed in PGCs and *Rhox6* has been found to be essential for PGCLC differentiation (Li et al., 2023b, Magnúsdóttir et al., 2013). We therefor wanted to see if *Parp1* deletion might impact *Rhox2, Rhox6* and *Rhox9* expression during PGCLC development. We performed qPCR analysis targeting *Rhox2, Rhox6* and *Rhox9* in mESCs, EpiLCs and PGCLCs day 2, day 4 and day 6 (Figure 5D). We found *Rhox2*, *Rhox9* and *Rhox6* expression elevated in the *Parp1^-/-^* day 6 PGCLCs, especially *Rhox2* and *Rhox6* which was significantly higher expressed when compared to *Parp1^wt/wt^* (p=0.001 and p=0.026, respectively, Wilcoxon test).

Proteomic and expression analyses revealed an upregulation of *Rhox* genes, specifically *Rhox2a/e, Rhox6* and *Rhox9*, in *Parp1^-/-^* cells. These findings suggest that PARP1 plays a role in inhibiting *Rhox* expression more broadly. Together with the proteomic analysis, the RNA-expression analysis suggests that PARP1 may downregulate *Rhox* gene expression, thereby increasing the likelihood of cells differentiating to PGCLCs in the absence of PARP1.

## Discussion

In this study, we identified that genetic variants in *PARP1* affects age at natural menopause and explored the role of PARP1 in early germ cell development using an *in vitro* model to generate primordial germ cell-like cells (PGCLCs) from mouse embryonic stem cells (mESCs). Our findings provide new insights into how PARP1 influences early germ cell formation and demonstrated its relevance to humans using a population genetics approach.

PARP1^V762A/V762A^ was associated with a significant reduction in day 6 PGCLC populations, representing the in vitro counterparts of early germ cells at approximately embryonic day 7 in mice (von Meyenn et al., 2016). The mouse embryonic stem cells (mESCs) carrying PARP1^V762A/V762A^ exhibited a lower total cellular NAD⁺ concentration compared to *Parp1^wt/wt^* cells. Given that NAD⁺ serves as a substrate for PARP1-mediated PARylation (Zong et al., 2022), this result suggests hyperactivation of PARP1^V762A/V762A^. Consistent with this, we observed increased levels of DNA-bound PARP1 in the *Parp1^V762A/V762A^*, which could indicate a prolonged chromatin retention or a higher amount of DNA bound chromatin, referred to as trapped PARP1. This is supported by our observations in *Xrcc1^-/-^* cells, which exhibit similar patterns of trapped PARP1 (Demin et al., 2021). Trapped PARP1 is known to be cytotoxic, consistently we also found upregulated apoptosis in the PGCLCs, especially on day four (Demin et al., 2021).

The variant investigated in this study (V762A), is located in the helical domain (HD) of the protein. PARP1 activation is tightly regulated by conformational (allosteric) changes in this HD. Upon binding to DNA lesions, the HD domain undergoes conformational shifts that are essential for initiating catalytic PARylation activity (Dawicki-McKenna et al., 2015). This highlights the importance of the HD domain in modulating PARP1 function and activity. PARP1 inhibitors provide further insight into HD domain function (Zandarashvili et al., 2020). Type I inhibitors destabilize the HD domain, leading to prolonged DNA retention of PARP1 in the DNA. In contrast, other inhibitors stabilize the HD domain, promoting faster release from DNA lesions (Zandarashvili et al., 2020). Furthermore, changes in the HD domain can differentially affect PARP1 activity, potentially leading to either hyperactivation or suppression of its enzymatic function (Dawicki-McKenna et al., 2015). Our data suggests that V762A could potentially have a destabilizing effect on the HD domain, leading to a prolonged chromatin retention and possible hyperactivation.

Cellular NAD⁺ levels may serve as a metabolic sensor influencing cell fate decisions. As metabolism plays a crucial role in differentiation, fluctuations in NAD⁺ availability could impact lineage specification (Tischler et al., 2019). In the context of PGCLC (primordial germ cell-like cell) formation, NAD⁺ depletion, such as that observed in certain PARP1 variants, may impair PARylation activity and disrupt key signaling pathways, ultimately leading to reduced PGCLC differentiation efficiency. This suggests that NAD⁺ availability could be a critical determinant in germ cell lineage commitment.

We hypothesize that the diminished pool of early germ cells could result in a smaller ovarian reserve, accelerating its depletion and contributing to an earlier onset of menopause, as observed in association with the *Parp1^V762A/V762A^*variant (Figure 6). This connection between germ cell population dynamics and reproductive lifespan warrants further investigation, as it could be a potential genetic marker for earlier onset of menopause for women.

**Figure 6:**
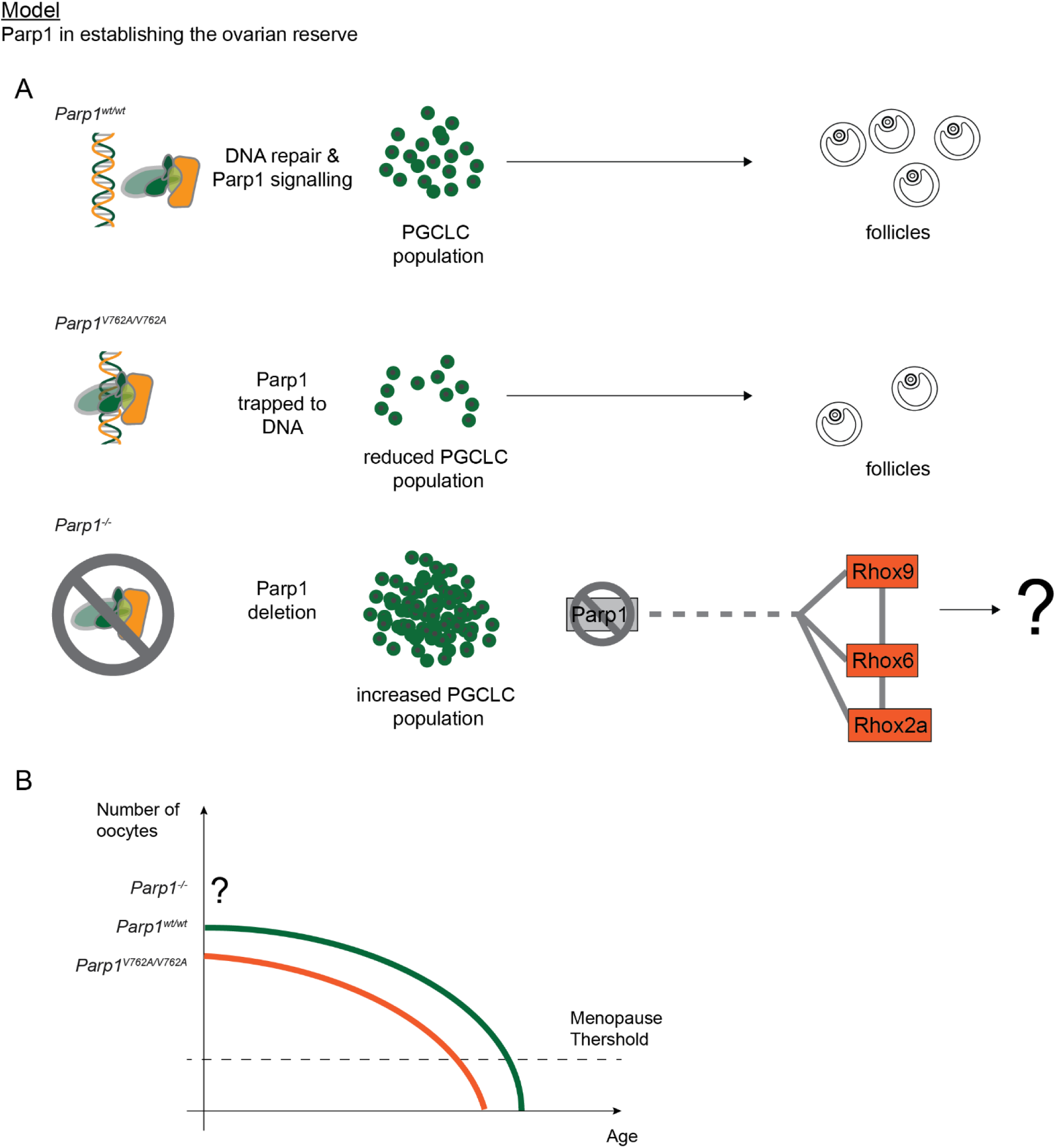
Summary of study and model of Parp1 in gametogenesis. **(A) Summary of Results:** *Parp1^V762A/V762A^* mouse embryonic stem cells exhibit a reduced number of primordial germ cell-like cells (PGCLCs), whereas the PGCLC population is upregulated *Parp1^-/-^* cells. **(B) Hypothesis on the role of Parp1 in ovarian reserve establishment and its correlation with ANM:** *Parp1^V762A/V762A^* displays a decreased number of primordial germ cells, potentially leading to an accelerated depletion of follicles throughout reproductive life and an earlier onset of menopause. In contrast, *Parp1^-/-^* cells, exhibit an increased PGCLC population. However, further studies are required to determine whether this increase translates to a higher ovarian reserve, as the long-term maintenance and function of these cells need to be further examined.

In contrast to the *Parp1^V762A/V762A^* cells, *Parp1^-/-^*cells displayed a significant increase in PGCLC populations compared to *Parp1^wt/wt^*, suggesting enhanced germ cell formation. This expansion was not accompanied by significant changes in proliferation rates, nor were there differences in the expression of key germ and stem cell markers, indicating that the germ cell identity of PGCLCs was maintained. Previous studies have shown that parthanatos, Parp1-induced cell death, acts as a mechanism to eliminate primordial germ cells (PGCs) in *Drosophila* that have migrated away from their intended location (Tarayrah-Ibraheim et al., 2021). While this mechanism could potentially explain the increased PGCLC numbers observed in our study, we found no significant differences in apoptotic markers. In fact, there was a slight increase in apoptosis on day 6 of the protocol. Notably, the phenotype observed in *Parp1^-/-^* cells is distinct from that of *Xrcc1^-/-^* cells, excluding base excision repair (BER) as the underlying mechanism.

One plausible explanation is a potential difference in transcriptional regulation or chromatin formation, which could influence PGCLC differentiation. To this end, we studied the proteome and RNA-expression of *Parp1^-/-^* and *Parp1^wt/wt^* cells. Interestingly, the proteomes showed minimal differences, however, RHOX2a, a protein implicated in early fetal ovary and testis development (Daggag et al., 2008) was significantly upregulated in *Parp1^-/-^* cells. Quantitative real-time PCR analysis further revealed increased expression of *Rhox6* and *Rhox9*, known to be critical for PGC formation (Li et al., 2023b, Magnúsdóttir et al., 2013).

We propose that PARP1 deletion may lead to the earlier expression of *Rhox* genes. This hypothesis is supported by the presence of elevated *Rhox* gene expression in the steady state (mESCs), suggesting that *Parp1^-/-^* cells may be primed toward PGC fate. However, *Parp1^-/-^* cells showed a slight increase in apoptosis by day 6 of differentiation, this suggests that while they may be more efficient in initiating germ cell development, they could encounter challenges at later stages due to the absence of *Parp1*.

Future research should explore these findings in vivo using knockout models to validate the role of *Parp1* in germ cell development. Additionally, studies should focus on the transcriptional and epigenetic mechanisms underpinning *Rhox* gene regulation and the connection to *Parp1*, as these may provide critical insights into the broader impact of *Parp1* on germ cell biology and reproductive lifespan.

---

**Figure S1:**
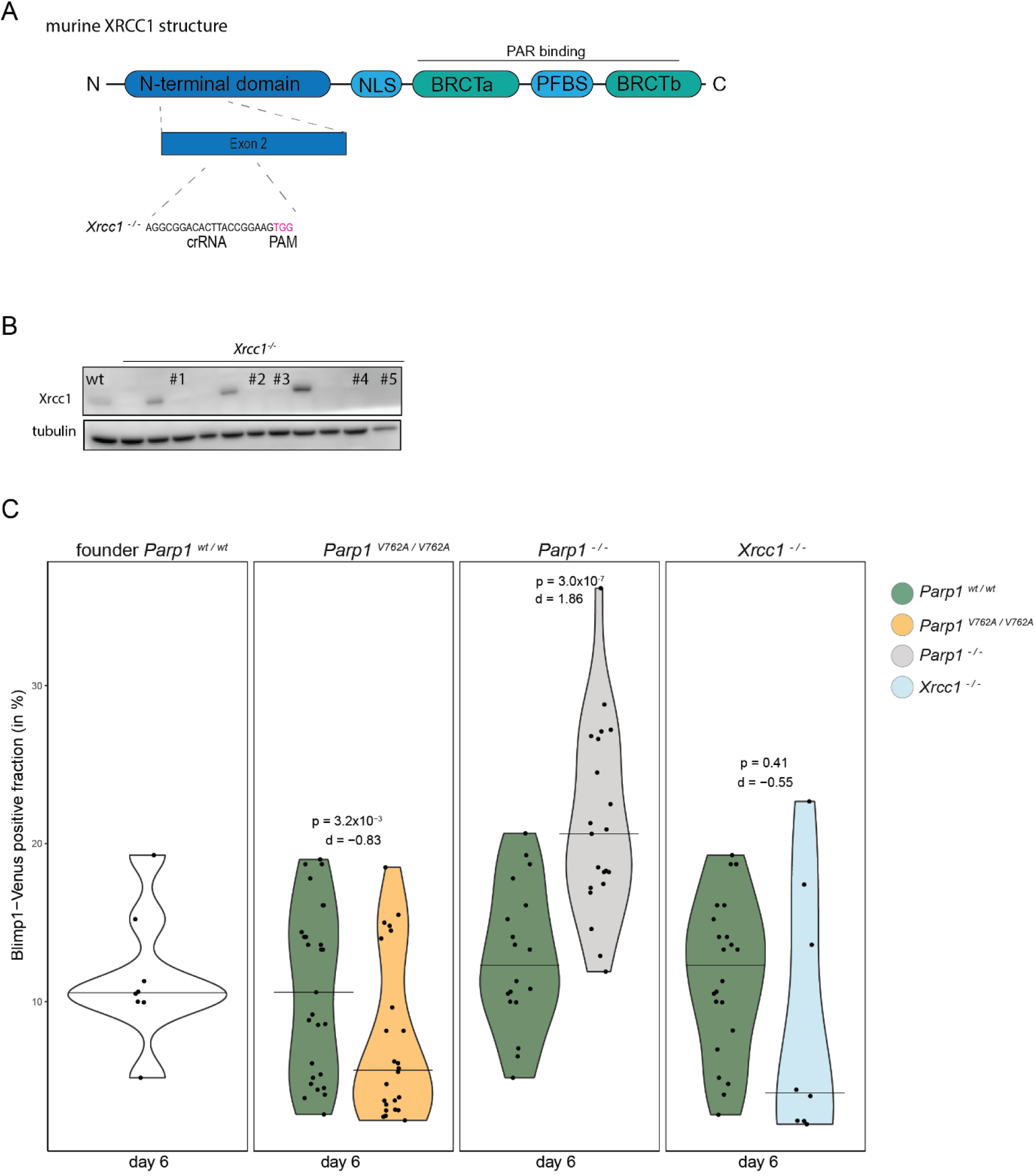
*Xrcc1 ^-/-^* cells do not replicate *Parp1* phenotype in PGCLCs. **(A)** schematic overview of Xrcc1 protein (mouse), with guide sequence for deletion **(B)** Western blot displaying tubulin and XRCC1 (70 kDa) protein and tubulin (55 kDa) for the *Xrcc1^-/-^* clones. Successful deletion of *Xrcc1* gene indicated by the absence of expected band **(C)** Violin plot displaying the percentage (%) of Blimp1-Venus positive cells on day 6, for the founder (not CRISPRed cells), the *Parp1^wt/wt^*that did not show PARP1 deletion after CRISPR as a control, the *Parp1^V762/V762A^*, *Parp1^-/-^* (from Figure 2) and *Xrcc1^-/-^.* Blimp1-Venus-positive PGCLC populations were quantified via FACS. Statistical significance was assessed using the non-parametric Wilcoxon test with Bonferroni adjustment. Effect sizes (Cohen’s d) are indicated.

**Figure S2:**
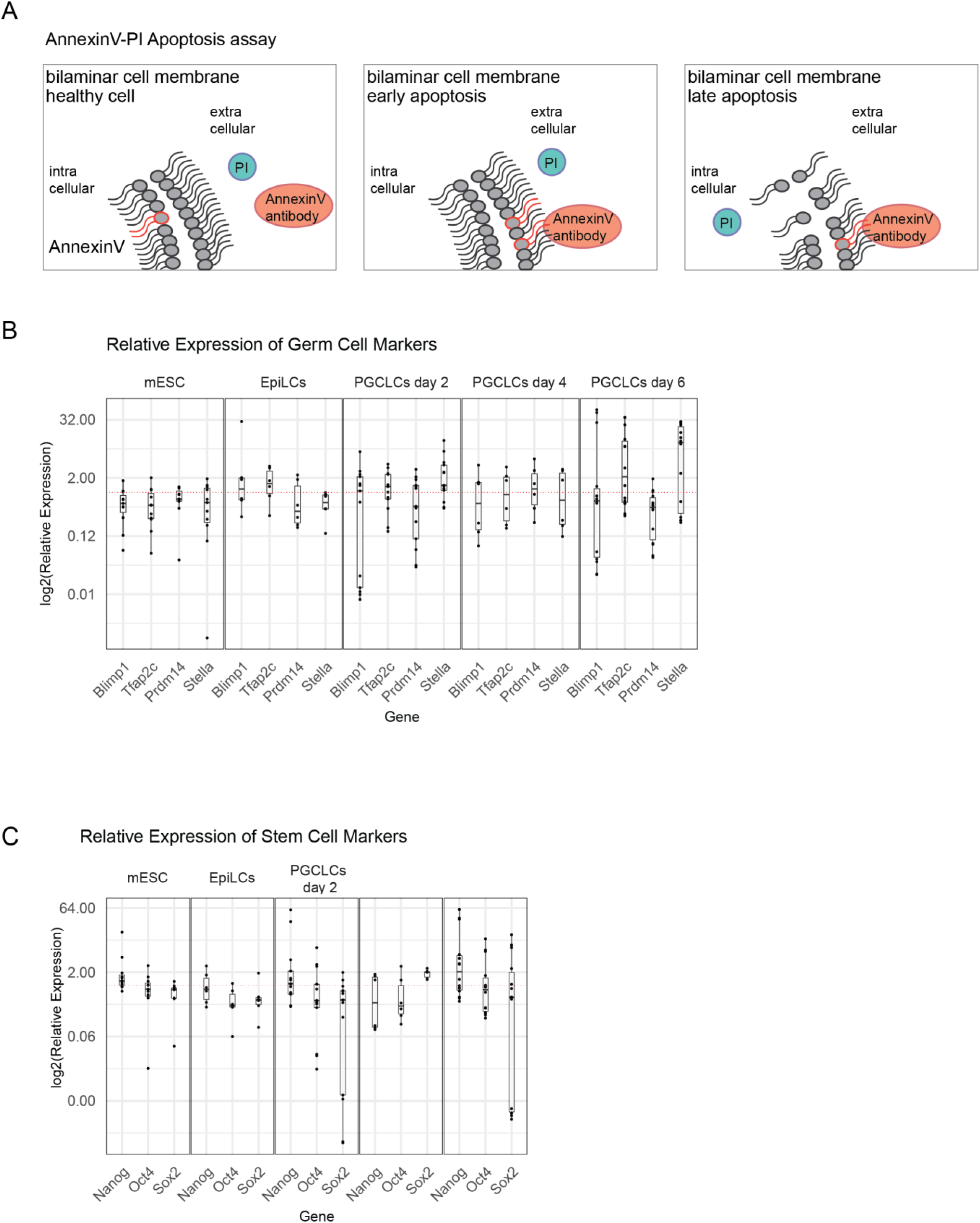
Schematic view of apoptosis assay and qPCR data from *Parp1^-/-^*. **(A)** Schematic representation of the apoptosis assay. In healthy cells, Annexin V remains associated with phosphatidylserine (PS) on the inner plasma membrane. During early apoptosis, PS is externalized to the outer membrane, where AnnexinV antibody can bind. In late-stage apoptosis, membrane integrity is lost, allowing propidium iodide (PI) to penetrate the cell and intercalate with DNA, indicating loss of membrane permeability (Vermes et al., 1995). **(B)** Boxplot depicting the quantitative PCR (qPCR) analysis of core stem cell markers (Oct4, Sox2, Nanog) in mESCs, EpiLCs, and FACS-sorted Blimp1-Venus-positive cells during PGCLC differentiation on days 2, 4, and 6. Data analyzed using the ΔΔCT method normalized to GAPDH and Parp1^wt/wt^ cells; statistical significance assessed by the non-parametric Wilcoxon test. **(C)** Boxplot of qPCR analysis of core germ cell markers (Blimp1, Stella, Prdm14, Tfap2C) in mESCs, EpiLCs, and FACS-sorted Blimp1-Venus-positive cells during PGCLC differentiation on days 2, 4, and 6. Data analyzed using the ΔΔCT method; statistical significance assessed by non-parametric Wilcoxon test.

## Methods

### Amino acid alignment

Alignment of murine and human PARP1 was performed using T-Coffee *(Notredame et al., 2000)*.

### mESC cell line description

H18 mouse embryonic stems cell line harboring a venus-Blimp1 and ECFP-Stella reporter were kindly provided by Takashi Yoshino, Department of Stem Cell Biology and Medicine, Graduate School of Medical Sciences, Kyushu University, Japan (Yoshino et al., 2021).

### mESC cell culture

mESCs were cultured in 2i media (SFES with the addition of 2mM L-Glutamine (#14190-094, ThermoFisher Scientific), 0.001 mM PD0325901 (#1036, Sellckchem), 0.00126% monothioglycerol (#M6145, Sigma), 1×10^3^ units/ml LIF (#ESG1107, Sigma) and 0.003 mM CHIR99021 (#1263, Sellckchem)) on Ploy-L-orthinine (0.01%, (#P3655, Sigma)) and laminin (300ng/ml, (#10152421, ThermoFisher Scientific) coated 6 well plates (#CLS3516, Corning Costar). Colonies were passaged by dissociating with TypLE Express Enzyme (#12604013, Gibco).

### PGCLC induction

PGCLC were differentiated as described by Hayashi et al., 2011 (Hayashi et al., 2011b). Briefliey, EpiLCs were induced by plating 1×10^5^ mESCs in a 12 well plate (#CLS3513, Corning Costar) on human plasma fibronectin (16.7µg/ml; #FC010-5MG, Sigma Aldrich) in N2B27 medium containing 20 ng/ml Activin A (#120-14, Peprotech), 12 ng/ml bFGF (#13256-029, Invitrogen) and 1% KOSR (#10828028, ThermoFisher Scientific). Media was changed on day one of culture. At day 2 of culture PGCLCs were induced by adding 2×10^3^ EpiLCs per well in lipidure coated low binding 96 well plate (#10344311, ThermoFisher Scientific) with GK15 (GMEM (Invitrogen, Waltham, MA, USA) with 15% KOSR, 0.1 mM NEAA (#11140050, ThermoFisher Scientific), 1 mM sodium pyruvate (#P5280, Sigma), 0.1 mM L-glutamine, 0.1 mM 2-mercaptoethanol (#21985023, ThermoFisher Scientific), 100 units Penicillin-streptomycin solution (#15140122, ThermoFisher Scientific) with the addition of the cytokines BMP4 (500ng/ml; #Q53XC5,R&D Systems, Minneapolis, MN, USA) BMP8b (500ng/ml; #P34820, R&D Systems) LIF (1000 u/ml; #A35935, Invitrogen), SCF (100 ng/ml; #Q78ED8, R&D Systems), and EGF (50 ng/ml; #P01133, R&D Systems). After day 2, day 4 or day 6 of culture PGCLCs were analysed /collected via flow through cytometry.

### CRISPR/Cas9

Crispr/Cas9 was done according to manufactures instruction (Inegrated DNA Technologies, Coraville, IA, USA). Briefly, tracr:crRNA duplexes were formed by mixing equimolar amounts of Alt-R™ crRNA (designed with: Integrated DNA Technologies) and Alt-R tracrRNA ATTO550 (#1975827, Integrated DNA Technologies) in IDT Duplex Buffer (30 mM HEPES, pH 7.5, 100 mM potassium acetate; Integrated DNA Technologies), heating to 95 °C for 5 min and slowly cooling to room temperature for 5-10 minutes. Ribonucleoprotein (RNP) complexes were formed by combining equimolar amounts of tracr:crRNA duplexes with Cas9 nuclease (#1081058, Alt-R *S.p.* Cas9 Nuclease, Integrated DNA Technologies) and incubated at room temperature for 20 min.

Alt-R™ HDR Donor Oligos (Integrated DNA Technologies) used in this study were resuspended using IDTE pH 7.5 (Integrated DNA Technologies). Sequences of the HDR oligos and guides used in this study are listed in Table 1.

### Delivery via Neon Transfection

Cells were transfected using the Neon Transfection System (#MPK1096, Invitrogen, Waltham, MA, USA). Cells were resuspended in Reuspension Buffer R at 1.0×10^7^ cells /ml. 10 µl of cells suspension was mixed with Electroporation Enhancer (100µM) and 1µl of RNP complex (described above). Mixture was taken up in the neon pipette (#MPP100, Invitrogen) and inserted into neon tube. Cells were electroporated 3 times at 1400V for 10 ms. Cells were immediately transferred into a 12 well plate coated with Poly-L-Ornithin and laminin and let attached overnight. The next day cells were sorted for ATTO-555 on Sony SH800Z (Sony Biotechnology, San Jose, CA, USA) into 96 well plates.

### Sanger Sequencing

To check DNA sequence after CRSIPR/Cas9 Sanger Sequencing was implemented. DNA was isolated using Qiagen DNA easy kit (#69504, Qiagen, Venlo, Netherlands). Sequences of interest were amplified using polymerase chain reaction (Q5-Hot-Start; #M0494S, New England Biolabs, Ipswich, MA, USA) and PCR product was purified using QIAquick PCR purification kit (#28104, Qiagen). Purified PCR products were mixed with primer (10 µM) and send to eurofins for sanger sequencing. All primers used in this study can be found in table X1.

### Protein collection

Cells were collected in cold Ripa (#R0278, ThermoFisher Scientific) supplemented with 1x Proteinase Inhibitor (#11836170001, cOmplete, Roche, Basel, Switzerland) and Benzonase (1:1000, #R0862, ThermoFisher Scientific). Samples were boiled with 1x SDS sample loading buffer at 95C for 5 minutes and then stored at - 20C.

### Chromatin fractionation

Soluble and chromatin bound PARP1 was separated as described by Gatti et al., 2020 (Gatti et al., 2020). Cell pellets were collected in 1x PBS supplemented with 1x protease inhibitor cocktail (#11836170001, cOmplete, Roche, Basel, Switzerland). For chromatin-bound and soluble fractions, cells were resuspended in chromatin extraction buffer (10 mM Hepes pH 7.6, 3 mM MgCl2, 0.5% Triton X-100, 1 mM DTT) supplemented with 1x protease inhibitor cocktail. Cells were rotated for 30 min at room temperature and spun down at 1300 g for 10 min at 4°C. The soluble fraction (supernatant) was centrifuged at 16 000 x g for 5 min at 4°C, collected again and boiled 1x SDS sample loading buffer at 95C for 5 min. The pellet of the chromatin extraction step (chromatin-bound fraction) was resuspended in MNase buffer (M0247S, New England Biolabs) supplemented with 1x protease inhibitor cocktail and 10 U of MNase (M0247S, New England Biolabs) for every 5 million cells. Cells were incubated for 30 min at 37°C, boiled in 1x SDS-loading buffer for 5 min at 95, spun down at 16 000 x g for 5 min and the supernatant was collected for immunochemical assays.

### Immunoblotting

Proteins were separated by standard SDS-PAGE (#NP0323BOX, ThermoFisher Scientific) and transferred onto PVDF membranes (iBlot™ 2 Transfer Stacks, IB24002, ThermoFisher Scientific) using the iBlot 2 system (IB21001, ThermoFisher Scientific). Membranes were blocked with 5% BSA in PBS-T (PBS + 0.1% Tween-20) for one hour at room temperature and incubated with primary antibodies diluted in 5% BSA in PBS-T over night at 4°C. Membranes were then washed three times for 15 minutes with PBS-T and incubated with HRP-conjugated secondary antibodies diluted in 5% BSA in PBST for one hour at room temperature and washed again three times with PBS-T. Protein signals were detected using ECL™ Western Blotting Detection Reagent (Amersham™). Antibodies used in this study can be found in table X2.

### qPCR

RNA was isolated from cells using RNaesy Kit (#74104, Qiagen, Venlo, Netherlands) and cDNA was synthesized using SuperScript II Reverse Trancriptase (#18064022, ThermoFisher Scientific). cDNA was diluted to 1 ng/µl and used for qPCR analysis with Power SYBR Green PCR MasterMix (#4368706, ThermoFisher Scientific). All primers used in this study are listed in Table 1.

### Apoptosis Assay

Apoptosis assay was performed using the eBioscience™ Annexin V Apoptosis Detection Kits (#88-8007-74, ThermoFisher Scientific). Briefly, cells were collected and washed with PBS and 1x binding buffer. Cells were resuspended at 5×10^5^ cells in 100µl and 5µl of AnnexinV-APC antibody was added. After 10 minute incubation at room temperature cells were washed in 1x binding buffer and resuspende in 200µl binding buffer. 5µl PI was added and cells were analyzed using the Cytek Aurora spectral flow cytometer (Cytek).

### Cellular NAD+ assay

NAD+ levels of cells were determined using NAD/NADH cell-based kit (#600480, Cayman, Ann Arbor, MI, USA) according to manufacturer’s instruction for non-adherent cells. Cells were harvested and for each well 7.5×10^5^ cells were added.

### Cryostections and Staining

Day 6 embryoid bodies (EBs) were washed with PBS and fixed in 4% PFA at 4C for 1 hour. After fixation, EBs were washed with PBS and incubated in 30% sucrose in PBS at 4C overnight. EBs were embedded in OCT the next day and blocks were kept at -20C. Blocks were sectioned on cryostat (xxx) at 10 µm and slides were stored in -20C. For staining, slides were dried at room temperature for 30 min and rehydrated in PBS for 5 min 2 times. Slides were blocked in blocking solution (3% BSA in PBST (PBS + 0.1% tween20)) for an hour at room temperature in a humid chamber. Primary antibodies diluted in blocking solution was added and incubated overnight in a humid chamber at 4C. The following day slides were washed 3 timed 10 minutes in PBST and secondary antibodies diluted in blocking solution was added for 1 hour at room temperature. Afterwards slides were washed 3 times 10 minutes in PBST and DNA was stained using DAPI (10µg/ml) (1:1000 in PBST) for 10 minutes. Slides were mounted using vectorshield (x, abcam) and stored at 4C until imaging on microscope XY. All antibodies used in this study can be found in table X2.

### Proliferation Assay

Proliferation of PGCLCs was analyzed via CellTrace™ Far Red Cell Proliferation Kit (#C34564 ThermoFisher Scientific). Day 2 EpiLCs were labelled for 30 minutes using CellTrace™ (1 µM in PBS) and after 120 hours PGCLCs (Blimp1-venus positive cells) were analyzed for Cell Trace using the Cytek Aurora spectral flow cytometer (Cytek).

### Statistics

Statistical analyses and data visualization were performed using R (Version 4.1.1). The Shapiro-Wilk test was employed to assess normal distribution. Normally distributed datasets were analyzed using t-tests, while non-normal datasets were evaluated with the non-parametric Wilcoxon rank-sum test. To account for multiple comparisons, p-values were adjusted using the Bonferroni correction method. Effect size (d) was calculated using Coheńs d. Visualizations were created using the ggplot2 package.

### Proteomics sample preparation

PARP1 WT and KO mESC cells (approximately 100,000 cells per replicate) were thawed on ice and lysed in SDS lysis buffer (2% SDS, 150 mM NaCl, 50 mM Tris pH 8). Samples were boiled at 95 °C with vigorous shaking at 1400 RPM and subjected to sonication to shear DNA using a Sonic Dismembrator (Fisherbrand) set at 20% amplitude for 15 seconds. Samples were reduced and alkylated by adding tris(2-carboxyethyl)phosphine (TCEP) and chloroacetamide (CAA) to a final concentration of 5 mM, followed by incubation at room temperature (RT) for 1 hour. Protein aggregation capture (PAC) digestion was performed as described previously (Batth et al., 2019). Breifly, proteins were aggregated onto 25 µL magnetic beads (MagReSyn hydroxyl beads, cat# MR-HYX2L, ReSyn Biosciences, Ltd) using acetonitrile at a final concentration of 80%. Beads were washed extensively with 100% acetonitrile and then with 70% ethanol.The beads were resuspended in digestion buffer (50 mM Tris, pH 8.5) containing 250 ng trypsin (Promega, Sequencing Grade) and incubated at 37 °C for 3 hours. Digestion was stopped by adding trifluoroacetic acid (TFA) to a final concentration of 1%. The supernatant was collected in a separate tube, and the beads were washed with 1% TFA and combined with the supernatant.Peptides were purified using low-pH StageTips prepared in-house with four plugs of C18 material (Sigma-Aldrich, Empore SPE Disks, C18, 47 mm) per StageTip. StageTips were activated with 100 µL 100% methanol, equilibrated with 100 µL of 80% acetonitrile/0.1% formic acid, and washed twice with 100 µL of 0.1% formic acid. Acidified samples were loaded onto the StageTips, washed twice with 100 µL of 0.1% formic acid, and eluted with 60 µL of 25% acetonitrile/0.1% formic acid. Peptides were dried to completion using a SpeedVac at 60 °C, dissolved in 11 µL of 0.1% formic acid, and stored at -20 °C until mass spectrometry (MS) analysis.

### Mass spectrometry analysis

All samples were analyzed using an Orbitrap Astral mass spectrometer (Thermo) coupled with a Vanquish Neo UHPLC system (Thermo) for on-line reversed-phase liquid chromatography. The analytical emitter was a 20- cm analytical column with a 75-µm internal diameter, packed in-house with ReproSil-Pur 120 C18-AQ 1.9 µm beads (Dr. Maisch). Peptides were eluted using a gradient from buffer A (0.1% formic acid) to buffer B (80% acetonitrile/0.1% formic acid) at a flow rate of 250 nL/min. The gradient length was 25 minutes per sample, including ramp-up and wash-out, with an analytical gradient of ∼20 minutes ranging from 7% to 35% buffer B.

The analytical column was heated to 40 °C in a column oven, and ionization was achieved using a NanoSpray Flex™ NG ion source. Spray voltage was set to 2 kV, the ion transfer tube temperature to 275 °C, and the RF funnel level to 50%. Full MS scans were acquired over an m/z range of 300–1000 with a resolution of 240,000, a 50-ms maximum injection time, and a 500% AGC target. Data-independent acquisition was performed with a 4-Th isolation window, a scan range of 100–1500 m/z, and a fragmentation energy of 25% HCD. MS2 scans were acquired with a maximum injection time of 5 ms, a 200% AGC target, and up to 175 MS2 scans per survey scan.

### Analysis of raw data

Raw data files were processed using DIA-NN (version 1.8.1) (Demichev et al., 2020). Output files were filtered at a 1% false discovery rate (FDR) using run-specific q-values. Precursor and protein x sample matrices were further filtered at a 0.002 precursor- and protein-level FDR (--matrix-qvalue 0.002) for additional stringency. Matching between runs and smart profiling were enabled. A mouse-specific FASTA file was downloaded from UniProt on November 6, 2023, for database searches.

### Data analysis and statistics

Statistical analysis of MS data was conducted using Perseus software (version 1.6.14.0)(Tyanova et al., 2016). Raw intensity values were log2-transformed, and gene groups were filtered to retain rows where at least one group had 4/4 valid values. Missing values were imputed column-wise (downshift: 1.8; width: 0.3). Differential expression analysis was performed using two-sample t-tests with a significance threshold of 5% FDR, applying permutation-based FDR with a fudge factor (s0) of 0.5. Figures were prepared using the R statistical programming language (version 4.3.1, “Beagle Scouts”) with the “ggplot2” package (version 3.4.2). FACS data was analyzed using FlowJo (Version 10.8, BD Life Sciences, USA).

## Data availability

The mass spectrometry proteomics data have been deposited to the ProteomeXchange Consortium via the PRIDE (Perez-Riverol et al., 2022), partner repository with the dataset identifier **PXD063073**.

## Supplementary Tables

**Table 1:**
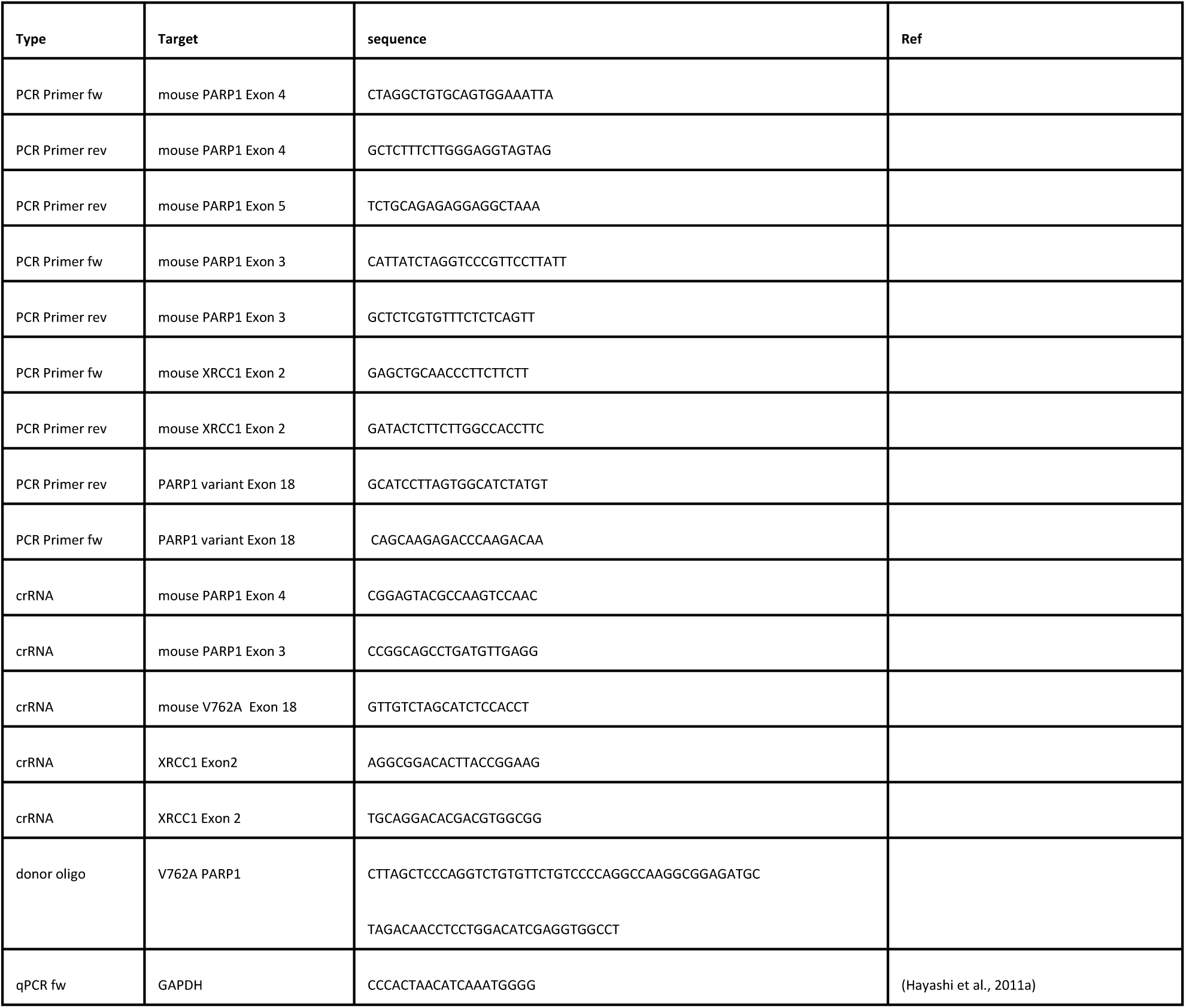
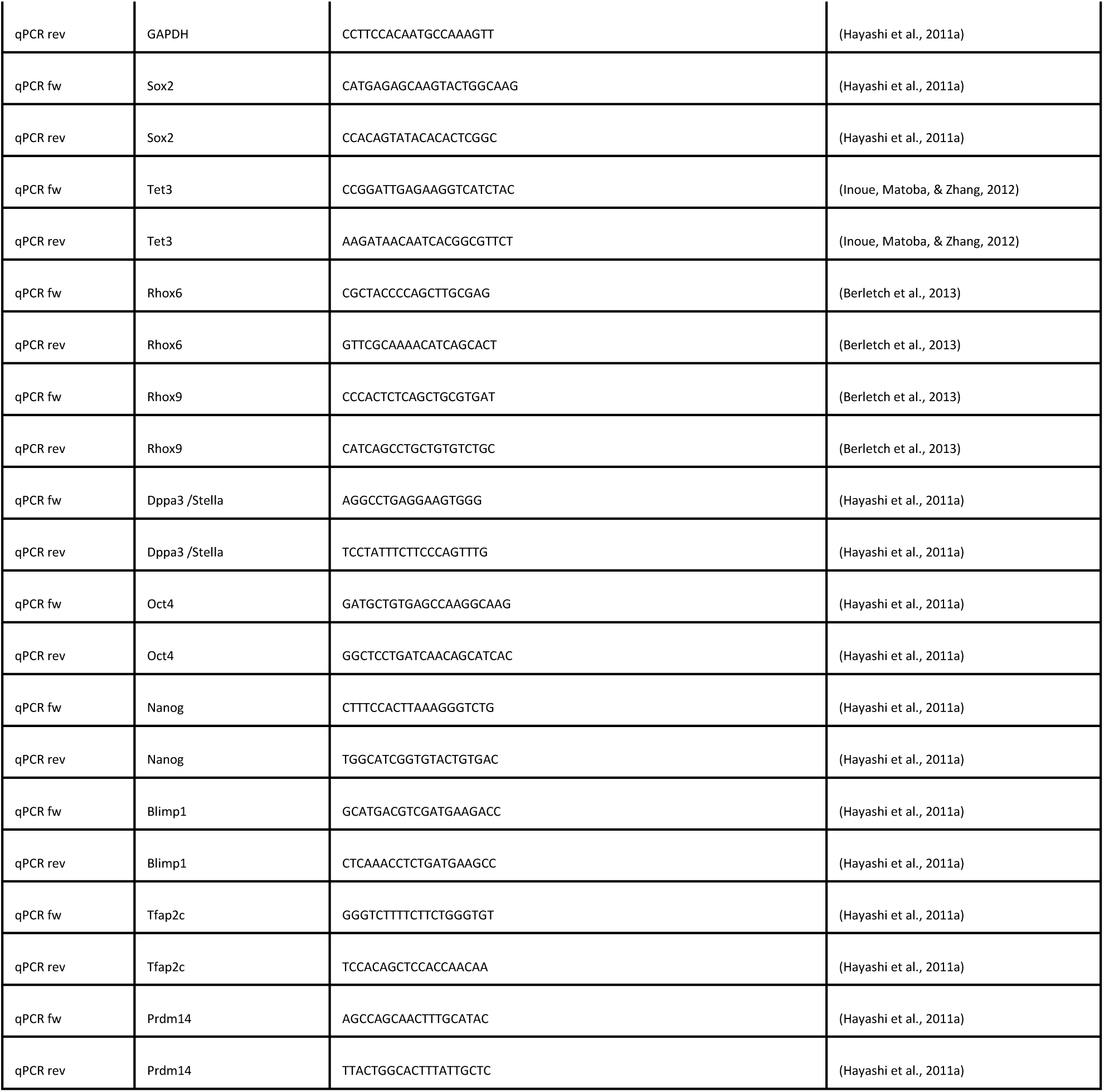
Oligos used in this study.

**Table 2:** antibodies used in this study

| 1st | company | # | [c] | 2nd | company | [c] |
| --- | --- | --- | --- | --- | --- | --- |
| PARP1 | abcam | ab191217 | 1 in 500 | Goat anti-rabbit HRP | Dako | 1 in 5000 |
| XRCC1 | abcam | ab134056 | 1 in 500 | Goat anti-rabbit HRP | Dako | 1 in 5000 |
| tubulin | Novus Biologicals | NB100-1639 | 1 in 2000 | Goat anti-rat HRP | invitrogen | 1 in 5000 |

**Table 3:** ANOVA results testing for factors influencing PGCLC fraction on day 6. PARP1: genotype status, either wild-type or V762A. ExpID: differences between experiments. clone: differences between clones. Degrees of Freedom (Df), Sum of Squares (Sum Sq), Mean Squares (Mean Sq), F-Values, P-Values (Pr(>F))

| group | Df | Sum Sq | Mean Sq | F value | Pr(>F) | factor |
| --- | --- | --- | --- | --- | --- | --- |
| V762A | 1 | 195,5818 | 195,5818 | 42,58274 | 1,39E-07 | genotype |
| V762A | 13 | 991,7193 | 76,2861 | 16,60927 | 1,9E-11 | ExpID |
| V762A | 16 | 412,9347 | 25,80842 | 5,619097 | 8,67E-06 | clone |
| V762A | 36 | 165,3474 | 4,592983 |  |  | Residuals |

## Acknowledgments

We would like to express my sincere gratitude to Elena Doncel and Amruta Shirkhande for their invaluable assistance with the Western Blot experiments. Their expertise and guidance were instrumental in optimizing the protocols. We are also grateful to the FACS core facility at reNew, ICMM, KU for providing access to the necessary equipment and resources. Additionally, we extend our appreciation to Gelo de la Cruz for his support with the FACS platform, including instrument operation, troubleshooting, and data acquisition. Their technical insights greatly contributed to the success of this study.

